# Predicting Methylation-Based Replication Timing from Whole Slide Images

**DOI:** 10.64898/2025.12.17.25342493

**Authors:** Alejandro Leyva, Abdul Rehman Ackbar, M. Khalid Khan Niazi

## Abstract

Replication timing is a costly but powerful tool for characterizing cellular mechanisms that underlie chromatin organization, cancer epigenetics, and genomic instability. Genome-wide replication timing profiles reflect the temporal order of DNA synthesis during S phase and are closely linked to chromatin accessibility, transcriptional activity, and proliferative state. Prior work has demonstrated a robust inverse relationship between replication timing and DNA methylation at the domain scale, enabling methylation-based proxies to approximate replication timing in large cohorts where direct experimental measurement is impractical.

Given the tight coupling between replication timing, chromatin structure, and cellular phenotype, we hypothesized that histologic morphology encodes information consistent with replication timing states. To test this hypothesis, we implemented Vision Transformer (ViT) architectures to predict replication timing proxies from whole-slide histopathology images. Patch-level embeddings were extracted using a pretrained ViT and aggregated through attention-based multiple instance learning to predict sample-level replication timing. In parallel, CellViT models were employed to perform cell-level prediction, enabling direct comparison between patch-based and cellular representations.

Across both modeling strategies, statistically significant correlations were observed between image-derived features and methylation-based replication timing proxies. Patch-level models achieved correlations of approximately 46%, while cell-level models consistently reached correlations exceeding 50% on the validation cohort. Prediction error remained stable across folds, with mean absolute error and mean squared error values ranging between 0.4 and 0.6.

These results demonstrate that replication timing–associated epigenomic states are reflected in tissue morphology and can be inferred using deep learning models applied to routine histopathology. This work establishes a feasible, noninvasive framework for replicosomic inference from whole-slide images and supports future efforts toward spatially resolved replication timing analysis and integrative modeling of replicative stress in cancer.

## 1 Introduction

Replicosomics is the study of the replication of genetic domains and genes during mitosis, particularly throughout the S phase of the cell cycle. Replication timing (RT) is used to characterize the temporal order of DNA synthesis and to probe chromatin stability, transcriptional potential, and proliferative state in differentiated and malignant cell lines (1,2). The rate at which specific genomic regions replicate early or late in S phase provides a functional index of chromatin accessibility and cellular state.

The field emerged around 2010 with the development of Repli seq, which enabled empirical measurement of replication timing in cultured cell lines (3). In this assay, bromodeoxyuridine is incorporated into newly synthesized DNA, followed by immunoprecipitation of early and late replicating fractions and sequencing to generate genome wide RT profiles (4). These profiles produce the canonical replication timing wave that distinguishes early euchromatic from late heterochromatic compartments. Although Repli seq provides high resolution ground truth, it is labor intensive, requires proliferating cells, and remains limited across tissue cohorts.

To address this limitation, multiple studies have demonstrated that replication timing can be inferred from DNA methylation data, particularly from Illumina 450K and EPIC arrays (5–7). These methylation based proxies exploit the inverse biological relationship between DNA methylation and replication timing at the domain scale. Late replicating domains, or partially methylated domains, are characteristically hypomethylated, whereas early replicating domains retain high CpG methylation (8–10). Most Repli seq studies define RT such that higher values correspond to earlier replication, producing a positive correlation between RT and methylation levels, typically r 0.3–0.6 (11). When RT is instead defined as replication lateness, the correlation becomes negative, reflecting progressive methylation loss in late replicating regions during cell division (12–14). While the sign of the correlation depends on the RT definition, the underlying biology is consistent: early replication corresponds to high methylation, and late replication corresponds to low methylation.

Because of this consistent relationship, methylation values can be statistically inverted to yield reliable domain level replication timing proxies. These proxies recapitulate the large scale organization of replication domains observed in Repli seq data and extend replication timing inference to tissue cohorts lacking direct measurements.

In parallel, advances in computational pathology have enabled tissue morphology to be encoded as high dimensional embeddings using Vision Transformers (ViTs). These models capture spatial texture, glandular architecture, and morphologic cues associated with cellular proliferation and differentiation (13, 15). Given the close coupling between replication timing, chromatin structure, and proliferative state, we hypothesize that slide level replication timing averages derived from methylation based proxies can be predicted directly from histologic image embeddings. In this study, we extract ViT patch embeddings from whole slide images and train regression models to predict mean replication timing, which we term the Replicosomic State, and evaluate its behavior in publicly available tumor datasets.

Clinically, replication timing reflects a transition from a static methylation landscape to a temporal replication program, where replicative stress is associated with activation of DNA damage response pathways such as ATR CHK1 and ATM CHK2 signaling (1–3). Elevated activity in these pathways has been linked to improved responses to replication associated chemotherapies, including platinum agents and topoisomerase inhibitors (4–6). Expression of regulators such as CHK1 and WEE1 also suggests sensitivity to epigenetic and S phase targeted therapies, including histone deacetylase inhibitors and DNA methyltransferase inhibitors, which act on replication coupled chromatin maintenance (7–9).

From a pharmacologic perspective, slide based replicosomics enables the conversion of methylation derived genomic states into replication based temporal features, allowing replicative stress to be spatially contextualized within tumor histology. While global patient level replication timing may be less informative than domain level patterns, spatially resolved replication stress can inform heterogeneity in cell cycle progression and DNA damage response activation. The ability of artificial intelligence models to infer replicative stress directly from hematoxylin and eosin images has not yet been systematically evaluated. We therefore hypothesize that Vision Transformer architectures can approximate replicative stress states from histopathology, providing a non invasive framework for replication phenotyping in clinical oncology.

In breast cancer, replication timing and replicative stress have demonstrated clear clinical relevance. Tumors deficient in BRCA1 or BRCA2 exhibit elevated replication stress and depend on ATR and CHK1 mediated stabilization of stalled replication forks (10). As a result, ATR inhibitors such as ceralasertib and elimusertib, as well as CHK1 inhibitors such as prexasertib, show selective activity in homologous recombination deficient and highly proliferative triple negative breast cancers (11–13). Replicative stress biomarkers have also been associated with improved response to platinum agents and PARP inhibitors including olaparib and talazoparib, which exploit defective fork repair (14–16). Epigenetic therapies that modulate methylation and chromatin structure, such as decitabine and vorinostat, are likewise being explored to resensitize resistant tumors through reprogramming of replication timing domains (17,18). Integrating replicosomic signatures with morphologic representations therefore provides a path from descriptive pathology toward functional inference of replication stress, bridging digital pathology with therapeutic stratification.

## 2 Materials and Methods

Bulk methylation data from .txt files and idat files were processed using the Bioconductor and limma packages in R version 4.4.0 on the Ohio Supercomputer Center(26). Samples were matched at the case level, with a typical 1:1 correspondence between methylation profiles and whole slide images, and occasional 1:2 methylation to slide mappings. The final cohort consisted of 900 unique cases. All model training and inference were performed on NVIDIA A100 GPUs.

Whole slide images for breast cancer were obtained from the TCGA-BRCA cohort. Bulk methylation data were processed independently and subsequently mapped to matched whole slide images. Replication timing proxies were generated at the domain level by segmenting partially methylated domains and smoothing beta values at a resolution of 200–400 kb. For evaluation, we report both domain level replication timing estimates and an aggregated genome wide mean replication timing score. Because the TCGA-BRCA methylation datasets provide .txt files containing beta value matrices, domain segmentation and smoothing could be applied directly.

Replication timing was inferred under the assumption that replication timing is inversely correlated with DNA methylation. This formulation follows prior work demonstrating that domain-scale DNA methylation levels encode the long-term replication schedule of the genome, with late-replicating domains undergoing progressive methylation loss and early-replicating domains retaining high methylation across cell divisions. For each genomic window *w*, replication timing was defined as

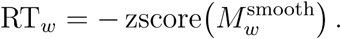

Smoothing was applied across genomic domains to normalize methylation values, as beta value matrices were preprocessed prior to analysis. The smoothed methylation value for each window was computed as

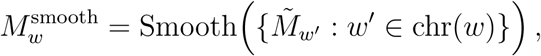

where the unsmoothed methylation value for each window was defined as

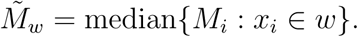

After smoothing, z scoring was applied across methylation domains such that regions with higher beta values correspond to earlier replication timing and regions with lower beta values correspond to later replication timing. Domain level replication timing was calculated as a length weighted average across constituent windows,

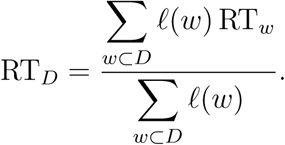

Here, *ℓ*(*w*) denotes the genomic length of window *w*. When window lengths varied, *ℓ*(*w*) was calculated as

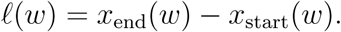

For uniformly sized bins, window length was fixed at

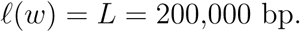

Sample level replication timing summaries were computed as both a length weighted mean replication timing score and the fraction of late replicating domains,

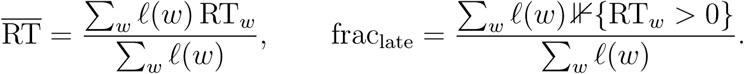

After sample level replication timing features were extracted, image based features were obtained using a baseline Vision Transformer (ViT). A ViT-B-16 architecture pretrained on ImageNet was used with frozen transformer blocks for feature extraction. Attention based multi instance learning was applied for patch weighting, followed by a multilayer perceptron regression head to predict replication timing proxies. Models were trained and evaluated using GroupKFold cross validation, with the prediction target defined as the sample level replication timing proxy. Statistical features derived from replication timing were retained for downstream comparison of sample variability but were not used as regression inputs. Model performance was evaluated using Pearson and Spearman correlation coefficients, as well as Mean Absolute Error (MAE) and Mean Squared Error (MSE). CellEcoNet was implemented to test predictions using 20x magnification UNIv2 Embeddings at the cellular level (27)

## 3 Results

Since RT is a continuous variable, the model used regression-based metrics such as correlation and MSE, rather than AUC. The results for the five fold classifications pass the pearson FDR test for statistical significance: The patch level ViT in Table 2 demonstrated moderate correlational strength and reasonable variance coverage across folds. While correlations did not consistently exceed 50%, this behavior is likely attributable to conservative batch sizes and a limited number of training epochs. Despite this, all folds exhibited statistically significant correlations and stable regression performance as measured by MSE and MAE. Performance was maximized in the third fold, where approximately 22% of the variance was explained. Across folds, MSE showed a consistent decline, whereas Pearson correlation peaked in fold 3. Spearman correlation was highest in fold 1 rather than fold 5, suggesting that monotonic relationships were more prominent in certain splits than strictly linear associations. MAE was included to assess the distribution of absolute errors and larger deviations, and closely tracked MSE across folds. Overall, the observed correlative strength is consistent with prior methylation based regression and prediction tasks, supporting the presence of epigenetically encoded signal within histologic morphology.

**Table 1:** Training configuration and hyperparameters used for RT mean regression.

| Parameter | Value |
| --- | --- |
| Epochs | 20 |
| Number of folds | 5 |
| Batch size | 1 |
| Maximum patches per case | 64 |
| Learning rate | $1 \times 10^{-4}$ |
| Weight decay | 0.05 |
| Frozen transformer blocks | 9 |
| Random seed | 42 |

**Table 2:** Performance metrics across validation folds for RT mean prediction.

| Fold | Pearson $r$ | $p$-value | Spearman $\rho$ | $p$-value | MAE | MSE | $R^2$ |
| --- | --- | --- | --- | --- | --- | --- | --- |
| Fold 1 | 0.2873 | $1.59 \times 10^{-4}$ | 0.3437 | $5.10 \times 10^{-6}$ | 0.5843 | 0.6184 | 0.0352 |
| Fold 2 | 0.3189 | $2.52 \times 10^{-5}$ | 0.1433 | 0.0639 | 0.5506 | 0.6904 | 0.0534 |
| Fold 3 | 0.4981 | $7.39 \times 10^{-12}$ | 0.2850 | $1.89 \times 10^{-4}$ | 0.4783 | 0.5124 | 0.2247 |
| Fold 4 | 0.4470 | $1.40 \times 10^{-9}$ | 0.2197 | 0.0043 | 0.5284 | 0.5190 | 0.1594 |
| Fold 5 | 0.4588 | $4.50 \times 10^{-10}$ | 0.1477 | 0.0568 | 0.4719 | 0.4643 | 0.1879 |

The cell based CellEcoNet model in Table 3,4, and 5 was evaluated using the original architecture reengineered with a regression head and trained for 100 epochs across five folds with explicit train, test, and validation splits [**?**]. Epochs corresponding to the lowest validation MSE were selected to represent training and test performance for each fold. Although evidence of overfitting was observed during training, both test and validation cohorts maintained strong Pearson and Spearman correlations. Across folds, MSE and MAE values were consistently between 0.4 and 0.6, and the majority of folds achieved Pearson correlation coefficients above 0.5. While variance explained was generally high, the test cohort of fold 5 exhibited low *R*^2^ despite a relatively high Pearson correlation, indicating stable ranking performance but reduced absolute variance capture. Spearman correlations remained moderately high across cohorts but were less consistent than Pearson correlations, reflecting weaker nonlinear monotonic structure.

**Table 3:**
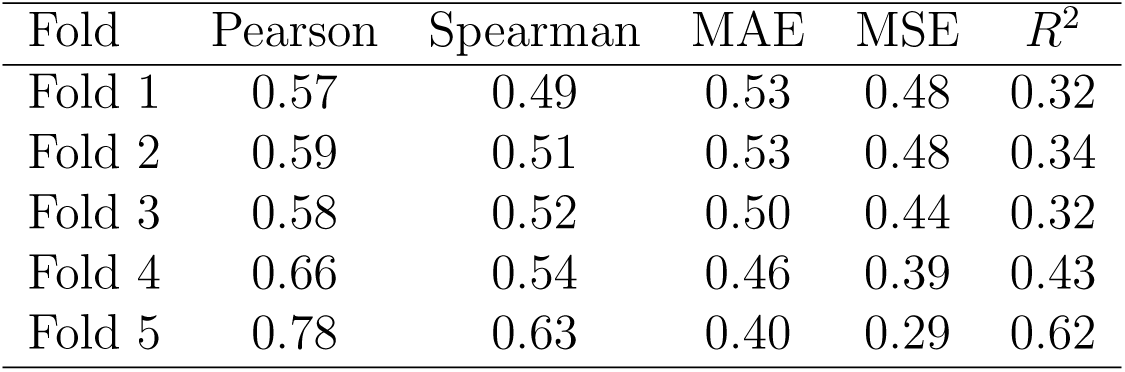
Training performance per fold CellEcoNet.

| Fold | Pearson | Spearman | MAE | MSE | $R^2$ |
| --- | --- | --- | --- | --- | --- |
| Fold 1 | 0.57 | 0.49 | 0.53 | 0.48 | 0.32 |
| Fold 2 | 0.59 | 0.51 | 0.53 | 0.48 | 0.34 |
| Fold 3 | 0.58 | 0.52 | 0.50 | 0.44 | 0.32 |
| Fold 4 | 0.66 | 0.54 | 0.46 | 0.39 | 0.43 |
| Fold 5 | 0.78 | 0.63 | 0.40 | 0.29 | 0.62 |

**Table 4:** Test performance per fold, CellEcoNet.

| Fold | Pearson | Spearman | MAE | MSE | $R^2$ |
| --- | --- | --- | --- | --- | --- |
| Fold 1 | 0.50 | 0.47 | 0.53 | 0.49 | 0.24 |
| Fold 2 | 0.65 | 0.54 | 0.45 | 0.39 | 0.40 |
| Fold 3 | 0.56 | 0.46 | 0.56 | 0.60 | 0.29 |
| Fold 4 | 0.51 | 0.40 | 0.62 | 0.56 | 0.15 |
| Fold 5 | 0.39 | 0.33 | 0.55 | 0.61 | 0.03 |

**Table 5:** Validation performance per fold, CellEcoNet.

| Fold | Pearson | Spearman | MAE | MSE | $R^2$ |
| --- | --- | --- | --- | --- | --- |
| Fold 1 | 0.67 | 0.54 | 0.44 | 0.35 | 0.45 |
| Fold 2 | 0.41 | 0.32 | 0.50 | 0.50 | 0.15 |
| Fold 3 | 0.55 | 0.47 | 0.49 | 0.45 | 0.30 |
| Fold 4 | 0.44 | 0.47 | 0.60 | 0.64 | 0.17 |
| Fold 5 | 0.63 | 0.43 | 0.40 | 0.32 | 0.262 |

Importantly, the improved performance of the cell based model relative to the patch level ViT likely reflects its ability to encode cellular neighborhoods and regional context. Replication stress is not confined to individual cells but emerges at the level of interacting cell populations, stromal architecture, and local microenvironmental structure. CellViT representations explicitly preserve spatial relationships between neighboring cells, enabling the model to capture coordinated stress responses, chromatin instability, and proliferative heterogeneity across cellular neighborhoods. This neighborhood level representation provides a more faithful substrate for predicting replication timing proxies, which themselves reflect domain scale and population level replication dynamics rather than isolated cellular events.

The learning curves for the patch level model, shown in Figure 1, demonstrate a stable decrease in loss over training, indicating no substantial overparameterization or underfitting. Figure 2 shows the distribution of predicted versus true replication timing values across validation folds. The bimodal concentration of low and high RT scores is biologically consistent, as mean domain level aggregates tend toward zero in samples with relatively preserved chromatin structure, whereas samples with widespread early replication across domains reflect chromatin instability and oncogenic transcriptional activation.

**Figure 1:**
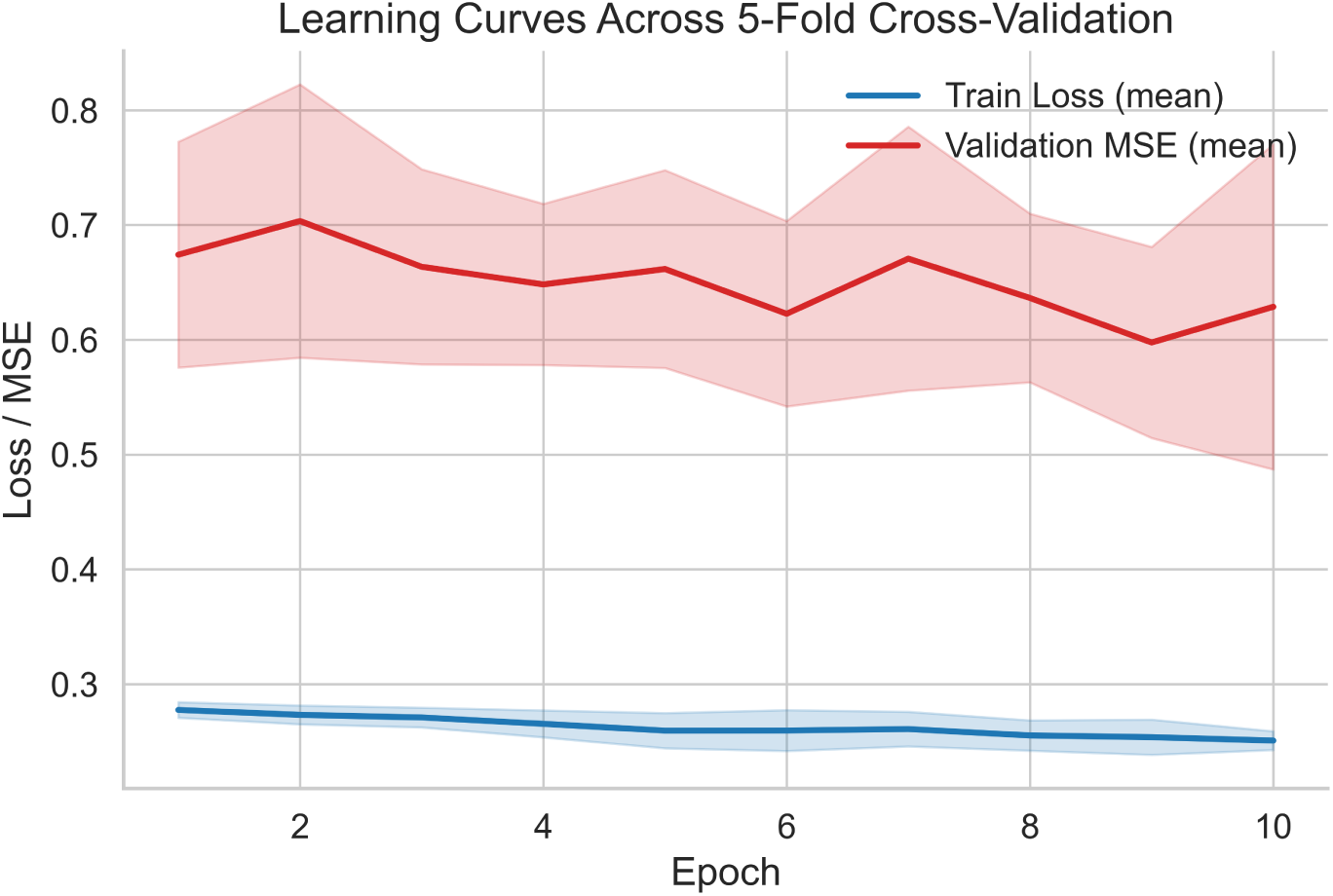
Learning dynamics across training. Mean (solid line) and ±1 SD (shaded) of training loss and validation MSE over epochs, aggregated across five folds.

**Figure 2:**
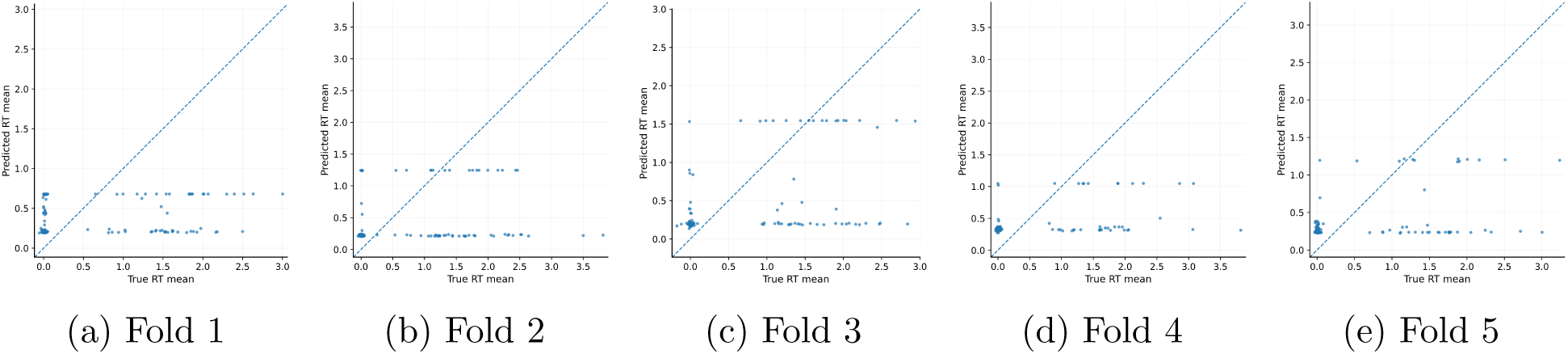
Predicted versus true RT mean across five validation folds. Each point represents a case, with the dashed line indicating *y* = *x*.

Prediction residuals are visualized in Figure 3. Improved alignment between predicted and expected values is evident in folds 3, 4, and 5, where case level deviations are reduced.

**Figure 3:**
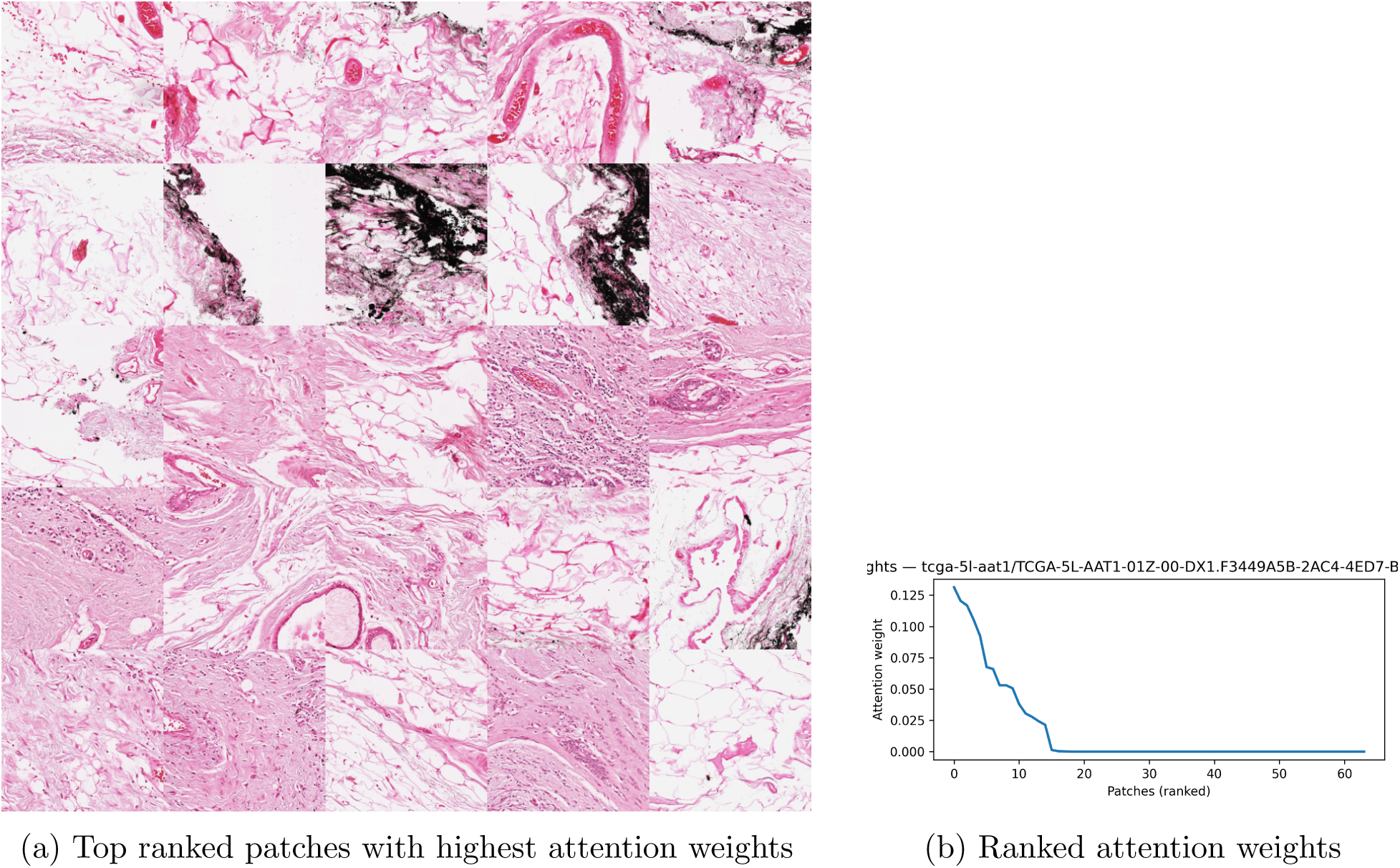
Representative attention distribution for the best qualitative case (tcga-4h-aaak). Attention is distributed across multiple informative patches rather than collapsing to a single focus, supporting spatially diffuse reasoning.

Attention maps from fold 3 demonstrate concentrated confidence over a restricted subset of patches, including adipose tissue, necrotic regions, and collagen rich stroma. These regions exhibit reduced methylation variance and lower nuclear density, consistent with late replication timing. In contrast, eosinophilic stroma with fibroblast activation and vascular regions show increased attention associated with earlier replication timing, reflecting replicative stress and chromatin instability at the regional level.

## 4 Discussion

The prediction model exhibits a higher MSE than traditional direct omics prediction models, which is expected given that replication timing is inferred indirectly through methylation based proxies rather than measured directly. Nonetheless, the observed correlations between morphology and replication timing are statistically significant, indicating that cellular phenotypes and tumor regions encode information consistent with early or late replication domains. Correlating replication timing with histologic morphology therefore provides a quantitative framework for assessing mechanistic changes in cellular phenotype, offering interpretability with respect to transcriptional activity, chromatin organization, and tumor progression (1,2).

Replication timing represents a higher order omics feature that integrates chromatin accessibility, proliferative state, and long range genomic organization, and can be combined with expression, mutation, and copy number data to support prognostic inference (3–5).

Importantly, even when derived from proxy level methylation data, replication timing retains a strong and reproducible correlative signal with tissue morphology, consistent with prior reports linking domain scale methylation patterns to replication dynamics across cancer types (6–8).

## 5 Limitations

This study relied on sample level averaged replication timing rather than domain resolved replication timing, which limits sensitivity to specific genomic regions with distinct transcriptional or oncogenic relevance. Patch level predictions were further constrained by conservative batching and a fixed 20 epoch training regime, with each epoch requiring approximately two hours due to embedding extraction, thereby limiting extensive hyperparameter exploration. Although the cell level model demonstrated improved performance, it also showed signs of overparameterization and late epoch overfitting.

The distribution of replication timing values was heavily skewed, reflecting transcriptional imbalance across samples, and may not fully represent broader patient populations. Additionally, because matched Repli seq data are not available for TCGA BRCA, direct verification of the methylation replication timing relationship within this cohort was not possible, despite substantial prior evidence demonstrating the robustness of this association across normal and malignant tissues (6–10). These limitations constrain the generalizability of the results until domain level replication timing and matched Repli seq validation data become available.

## 6 Conclusion

Replication timing and the ordering of early and late replicating domains are detectable within cellular phenotypes, with correlations exceeding 50 percent and explaining approximately 45 percent of variance across tissue samples and cellular neighborhoods. This establishes a feasible pathway for predicting methylation linked structural chromatin changes across cell populations and for inferring chromatin accessibility patterns associated with replicative stress. Clinically, replication timing derived from histologic images may approximate tumor state, oncogenic programs, treatment response, or disease progression, although further validation is required.

The use of methylation based replication timing proxies substantially reduces experimental burden relative to Repli seq and enables analysis across large public cohorts. Both patch level and cell level models indicate that attention is preferentially allocated to biologic structures associated with replicative stress and replication stability, including collagen rich stroma and regions of altered cellular density. Collectively, this work provides a structured view of how replication timing manifests across tissue morphology and supports future efforts toward domain resolved replication timing prediction and clinically actionable prognostic modeling.

## Data Availability

All data is available in TCGA and code is inrepo https://portal.gdc.cancer.gov/projects/TCGA-BRCA
https://github.com/Alejandro21236/ReplicationTiming

## 7 Ethics Statement

All datasets are public and anonymized.

## 8 Author Contributions

All work was performed by the first author, CellEcoNet was developed by the second author, with supervision by the third author.

## 9 Conflict of Interest

No conflicts of interest have been declared

## 10 Data Availability

TCGA BRCA methylation datasets and diagnostic slides are publically available and anonymized.

## Notes

### Competing Interest Statement

The authors have declared no competing interest.

### Funding Statement

No funding

### Author Declarations

TCGA BRCA is a publically available repository.https://portal.gdc.cancer.gov/projects/TCGA-BRCA All data is anonymized and informed consent was approved

